# Cross-scale prediction of glioblastoma MGMT methylation status based on deep learning combined with magnetic resonance images and pathology images

**DOI:** 10.1101/2025.05.07.25327185

**Authors:** Xusha Wu, Wei Wei, Yan Li, Menghang Ma, Zhenyuan Hu, Yongqiang Xu, Wenzhong Hu, Gang Chen, Rui Zhao, Xiaowei Kang, Hong Yin, Yibin Xi

## Abstract

**Background:** In glioblastoma (GBM), promoter methylation of the O^6^-methylguanine-DNA methyltransferase (MGMT) is associated with beneficial chemotherapy but has not been accurately evaluated based on radiological and pathological sections. To develop and validate an MRI and pathology image-based deep learning radiopathomics model for predicting MGMT promoter methylation in patients with GBM.

**Methods:** A retrospective collection of pathologically confirmed isocitrate dehydrogenase (IDH) wild-type GBM patients (n=207) from three centers was performed, all of whom underwent MRI scanning within 2 weeks prior to surgery. The pre-trained ResNet50 was used as the feature extractor. Features of 1024 dimensions were extracted from MRI and pathological images, respectively, and the features were screened for modeling. Then feature fusion was performed by calculating the normalized multimode MRI fusion features and pathological features, and prediction models of MGMT based on deep learning radiomics, pathomics, and radiopathomics (DLRM, DLPM, DLRPM) were constructed and applied to internal and external validation cohorts.

**Results:** In the training, internal and external validation cohorts, the DLRPM further improved the predictive performance, with a significantly better predictive performance than the DLRM and DLPM, with AUCs of 0.920 (95% CI 0.870–0.968), 0.854 (95% CI 0.702–1), and 0.840 (95% CI 0.625–1).

**Conclusion:** We developed and validated cross-scale radiology and pathology models for predicting MGMT methylation status, with DLRPM predicting the best performance, and this cross-scale approach paves the way for further research and clinical applications in the future.

## Introduction

Glioblastoma (GBM) is a highly aggressive form of cancer and is considered one of the deadliest cancers, with a poor prognosis for most patients. It is widely believed that MGMT promoter methylation status is a favorable prognostic factor in patients with GBM, being associated with a stronger response to alkylating agents such as temozolomide, a higher response to radiotherapy, and a longer survival time, with MGMT promoter methylation being present in approximately 45% of cases (Hegi et al., 2005). Therefore, selecting this group at the time of disease diagnosis will help achieve optimal treatment for responders and protect other patients from overtreatment-related toxicity.

Magnetic resonance imaging (MRI) is the imaging modality of choice for the diagnosis and monitoring of malignant tumors of the central nervous system (Guzman-De-Villoria et al., 2014). The development of MRI-based radiomics has provided additional information that is undetectable by the human eye to assist the traditional diagnostic radiologic workflow (Gillies et al., 2016). Current radiomic studies in the field of glioma have shown promising results in demonstrating correlations between MRI features and GBM molecular features and prognosis (Xi et al., 2018; Doniselli et al., 2023; Guan et al., 2023). However, conventional radiomics requires laborious and time-consuming precise tumor labeling, and preselection of morphological features limits the information that can be extracted from the images. In addition, recent studies have suggested that radiomics models alone may not be sufficient to accurately predict MGMT promoter methylation status in patients with gliomas prior to surgery (Brancato et al., 2022b; Doniselli et al., 2024).

It is well known that basic prognostic data is embedded in pathology images. Currently, a substantial body of research has successfully predicted tumor biomarkers using pathological slides (Niehues et al., 2023; El Nahhas et al., 2024; Volinsky-Fremond et al., 2024a; Wang et al., 2024). We assumed that the combination of macroscopic MRI and microscopic pathology images of tumors can more accurately reflect the biological information, and that the integration of the corresponding technologies will make it easier to break through the bottleneck of constructing cross-scale predictive models, and will be more relevant to clinical practice. In addition, pathology slides are more readily available in clinical practice for their affordability and low technical requirements compared to molecular testing. Thus, linking potential MRI features to pathological features could provide information across scales (Brancato et al., 2022a) and provide complementary sources of information about tumors in vivo and in vitro. In this study, we developed and validated the deep learning radiomics model (DLRM), deep learning pathomics model (DLPM), and deep learning radiopathomics model (DLRPM) for predicting MGMT expression in GBM patients based on preoperative multiparameter whole brain MRI images and postoperative digital pathology.

## Methods

### Patient Cohorts

We collected two independent patient cohorts from China. First, the primary cohort, including the training cohort and the internal validation cohort. This cohort comprised 182 Chinese patients from Xi’an People’s Hospital (Xi’an Fourth Hospital) (10 cases) and Xijing Hospital (172 cases), China, with pathologically confirmed GBM between September 2015 and September 2023. MGMT methylation levels were determined using pyrosequencing (the specific methods are as described in our previous publication), with a methylation rate greater than or equal to 10% interpreted as positive, otherwise negative (Xi et al., 2018). The inclusion criteria were (a) it conforms to the 2021 World Health Organization Classification of Central Nervous System Tumors and is classified as GBM (IDH-wildtype) based on histological type and molecular testing; (b) MRI scans available within 2 weeks before surgery; (c) pathological tissue slides (hematoxylin and eosin staining) stored as digital whole-slide images (WSIs). The exclusion criteria included (a) missing MRI data (incomplete T1WI, T2WI, or Gd-enhanced T1WI sequences) or insufficient quality or format conversion failure, (b) inability to determine MGMT status, (c) IDH mutant GBM, (d) blurred or unclear pathological images. Of the 182 patients in the primary cohort, 38 have been previously reported (Xi et al., 2018). The previous article discussed the prediction of MGMT using only traditional radiomics, which requires manual outlining of target areas, whereas in this study, we build on the previous work to break through the bottleneck of simple radiomics models and improve the prediction accuracy by jointly applying deep learning pathomics, i.e., we used the deep learning radiopathomics method and extract features for the prediction of MGMT with an aim to improve the prediction accuracy and generalization ability of the model.

Second, we collected a nonoverlapping cohort of 25 patients with GBM diagnosed at another Chinese center between August 2013 and June 2016 as a separate external validation cohort. The inclusion and exclusion criteria were identical to the primary cohort. Finally, 207 patients were included in this study.

### The Acquisition and Preprocessing of Imaging

The primary dataset was obtained using a 3.0T MR Siemens Magnetom Trio Tim with an 8-channel head coil and a 3.0T MR Siemens MAGNETOM Prisma with a 20-channel head coil. The head of the patient was fixed with a sponge cushion, and the patient was supine on the examination table. The scan sequences included axial T1WI, axial T2WI, and axial post-contrast T1. Sequence specific parameters are in the Supplementary Information. The external validation dataset also includes axial T1WI, axial T2WI, and axial post-contrast T1, all obtained on a 3.0T MR Siemens Tim Verio scanner with an 8-channel head coil.

For MRI preprocessing, we used the skull stripping algorithm HD-BET (Isensee et al., 2019) to remove skull and non brain tissue in MRI. The T1WI of each patient was aligned to the MNI 152 T1WI standard template using FSL-FLIRT (Jenkinson et al., 2002). We then aligned each patient’s T2WI and Gd-enhanced T1WI with T1WI, and trilinear interpolation was used during the alignment process to generate new images. For the pathology images, the WSI of the primary cohort was obtained by scanning biopsy tissue slices with 40× magnification (resolution: approximately 0.242 μm/pixel) and the external validation cohort was obtained by scanning with 20× magnification (resolution: approximately 0.485 μm/pixel). We used CLAM (Lu et al., 2021b) to segment out the tissue regions in WSI, and then tiled the 40× magnification WSI into multiple 512×512 pixel patches and the 20× magnification WSI into multiple 256×256 pixel patches to ensure that each patch has the the same receptive field.

### MGMT-Related Radiopathomics Features Extraction

Prior to feature extraction, we normalized the images to better fit the pre-trained model. Specifically, the resolution of all MRI images was adjusted to 224 × 224 × 224 and the MRIs were intensity normalized according to the mean and standard deviation of the brain tissue regions. The WSI Patches were adjusted to 224 × 224 and normalized according to the mean and variance of the ImageNet dataset. In this study, two types of pretrained models, the pre-trained 3D ResNet-50 (He et al., 2016), was used to extract the whole-brain deep learning radiomics (DLR) features of MRI and deep learning pathomics (DLP) features of WSI, respectively. The 1024-dimensional features output from global average pooling of the last convolutional layer of the third residual block of 3D ResNet-50 were used as the MRI sequence-level features, and the sequence-level features belonging to each patient were computed as the average to obtain the DLR features for each patient. The 2D ResNet-50 model was pretrained on the ImageNet (Deng et al., 2009) dataset. Similar to the extraction of MRI sequence-level features, the features extracted from pre-trained 2D ResNet-50 were used as WSI patch-level features, and the average value was calculated along the patch axis so that all patch-level features of each patient were fused into DLP features. Finally, a total of 1024-dimensional DLR features and DLP features were extracted in this study. The DLR and DLP features were concatenated to form a composite set of deep learning radiopathomics (DLRP) features for the patients.

### Model development

We randomly divided patients from the primary cohort into a training cohort and an internal validation cohort at the 8:2 ratio. Before modeling, in order to avoid redundant features affecting the generalization of the model, a coarse-to-fine strategy was used to select the optimal features related to the MGMT genotyping. Specifically, the Pearson correlation coefficient of the training cohort features and labels is first sorted from high to low, and the N-dimensional (*N* ∈{32,64, 96,128,160,192}) features most closely related to MGMT are selected as the preliminary selection results. The least absolute shrinkage and selection operator (LASSO) is used to analyze the nonlinear relationship between the initially retained features and labels. Finally, an MGMT classification model is built based on high-quality features with non-zero weights and the logistic regression method. DLRM, DLPM, and DLRPM were constructed using DLR features, DLP features, and DLRP features, respectively, to predict the MGMT methylation status of GBM patients (Figure 1). This study employs Class Activation Mapping (CAM) (Zhou et al., 2016) to visualize the features extracted by the 3D ResNet-50 and 2D ResNet-50 networks (Figure 2). This approach helps us understand the image regions that the networks focus on, and through meticulous manual review, we further validate whether these features are associated with the tumor.

**Figure 1.**
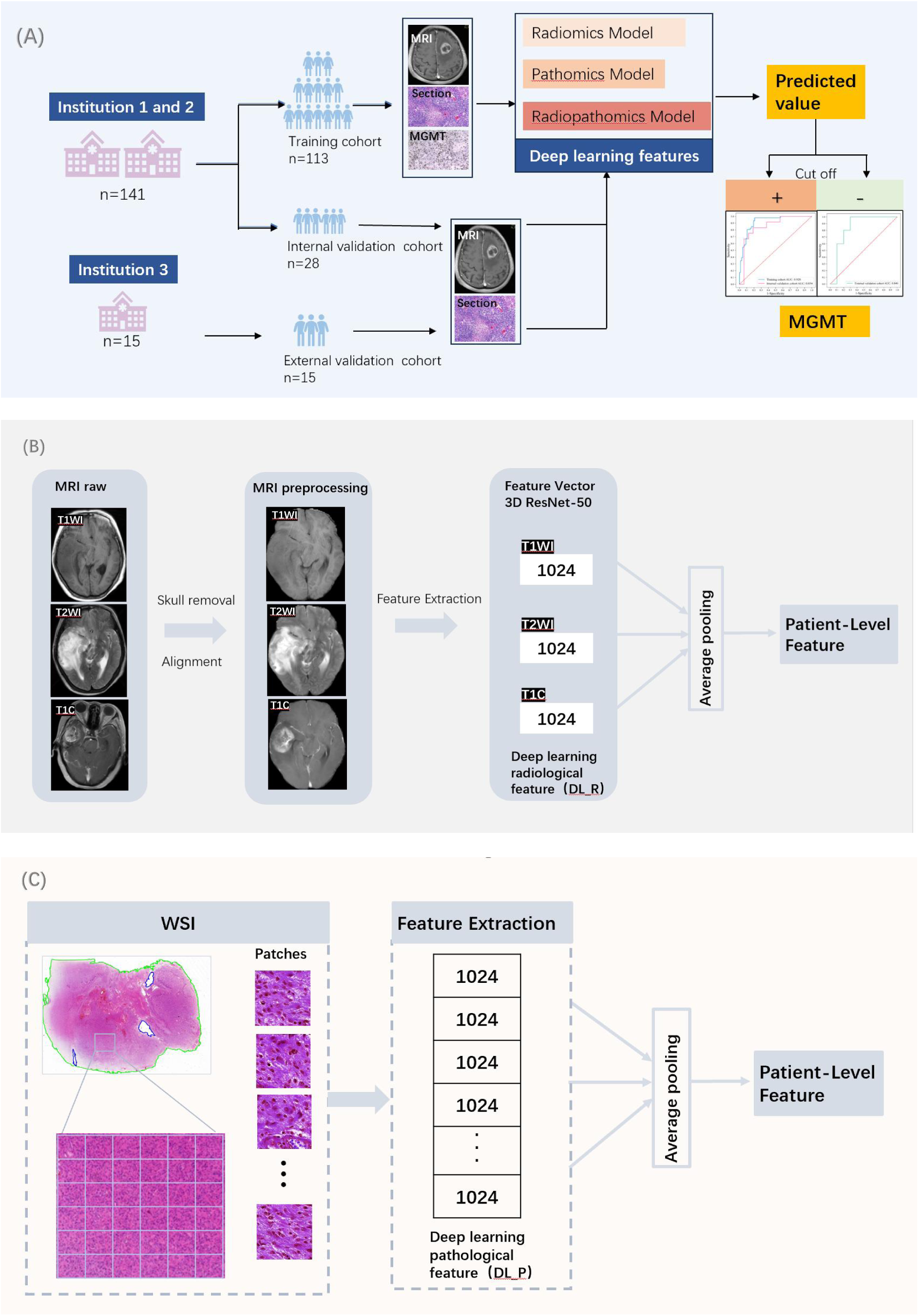
Overview of the workflow. (A), the training and validation process of the deep learning radiopathomics model is shown. (B), demonstrates the deep learning radiomics feature extraction process. (C), demonstrates the deep learning pathomics feature extraction process.

**Figure 2.**
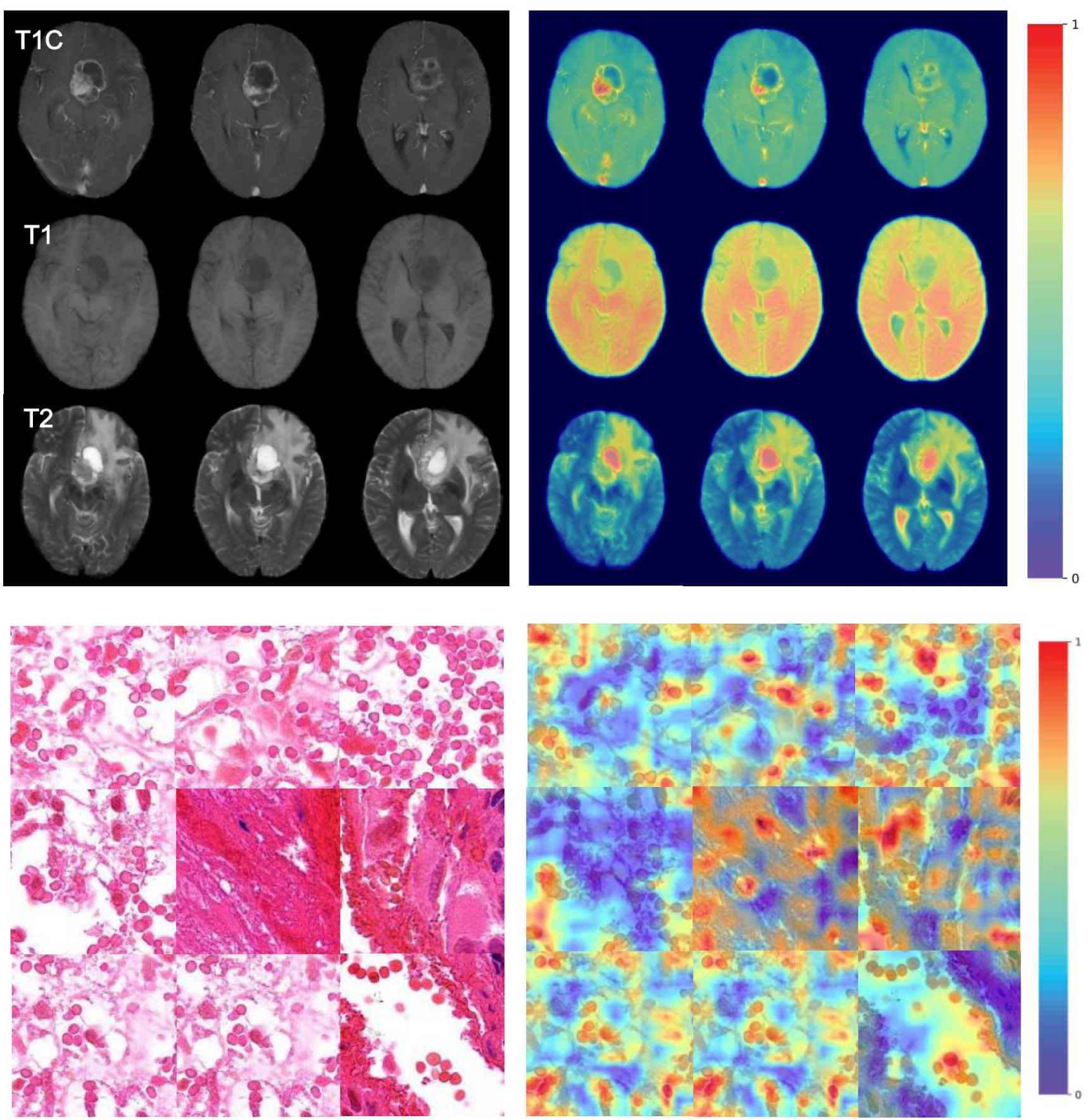
Class Activation Mapping (CAM) was utilized to visualize the features extracted by both 3D ResNet-50 and 2D ResNet-50 networks. Warm color tones indicate higher attention levels, while cool color tones represent lower attention.

### Visual MRI Feature Evaluation

Visual MRI features of GBM patients were collected, including lesion location, cystic necrosis, strengthening mode, subventricular zone affected, edema, and number of lesions. Based on a previous study (Gatto et al., 2022), a morphological assessment of the tumor was performed: (a) The location of the lesion was determined by the lobe in which the center of the lesion was located. When multiple lesions were present, the lobe with the largest lesion was used as the criterion and was divided into 5 groups: left frontal lobe, right frontal lobe, left temporal lobe, right temporal lobe, and other brain regions. (b) The strengthening modes were categorized into circular enhancement and non-circular enhancement, where non-circular enhancement included non-enhancement and other enhancement modes except circular enhancement. (c) Cystic necrosis area was defined as a nonenhanced area showing high signal on T2WI. (d) Whether the subventricular zone was affected. (e) Peri-lesion edema greater than or up to 1/3 of the lesion was considered serious. (f) Single lesion or multiple lesions. Patient characteristics were shown in Table 1.

**Table 1:**
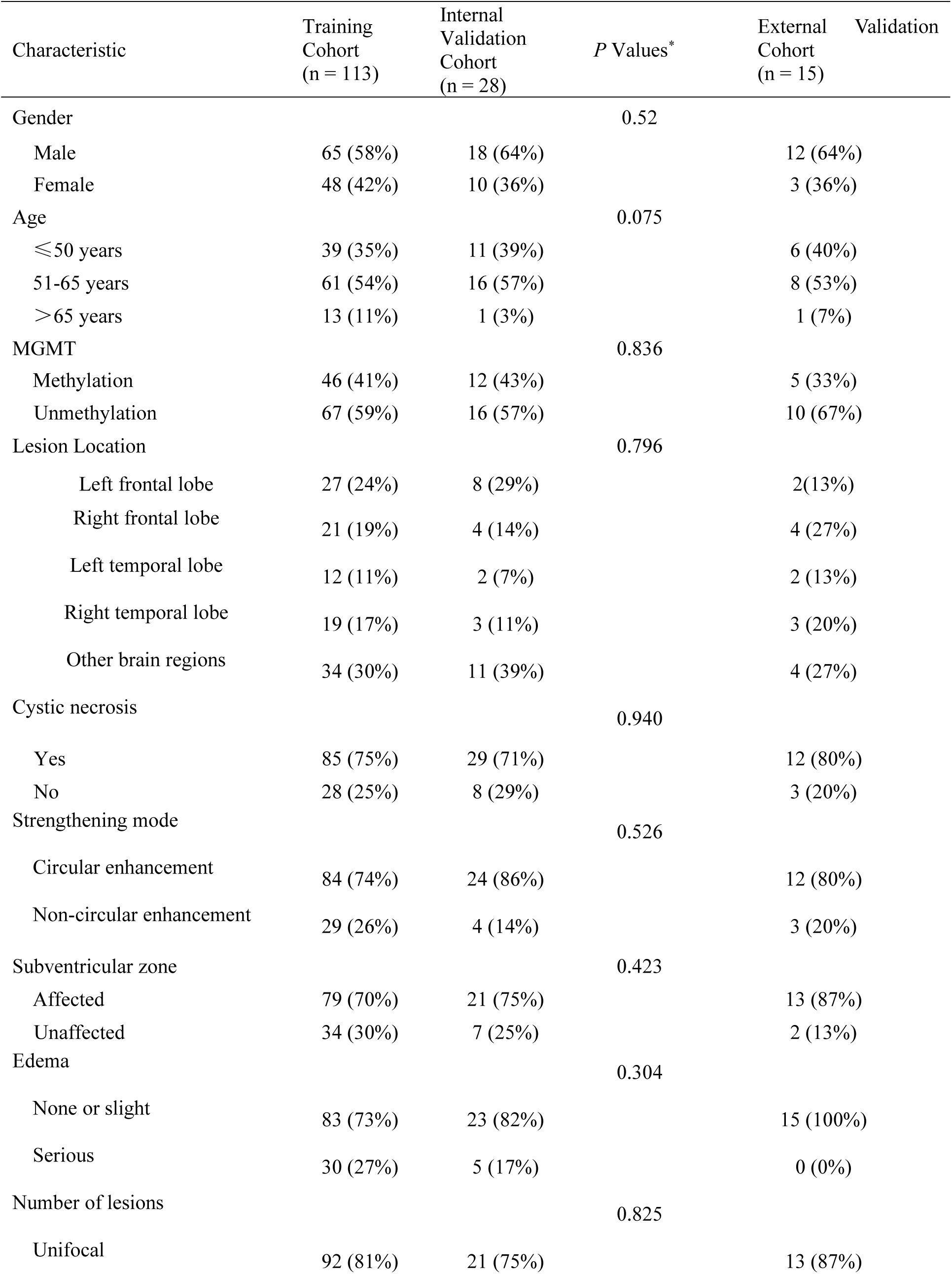
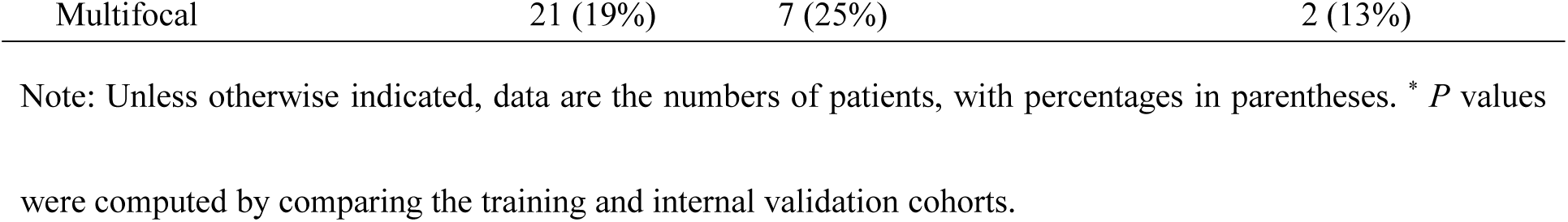
Patient Characteristics in the Training, Internal Validation, and External Validation Cohorts.

### Statistical analysis

Statistical analysis was performed using SPSS software (version 25.0). U-test was used for numerical variables and chi-square test for categorical variables to analyze demographic information. Non-parametric tests were used for clinical information assessment. ROC analysis was used to assess predictive performance (R software, version 4.2.2). Calibration curves were used to represent the gap between predicted and actual values (R software, version 4.2.2). Decision curves analysis (DCA) was used to compare the net benefits of DLRM, DLPM, and DLRPM (R software, version 4.2.2). Correlation analysis between model predicted values and visualized MRI features. The statistical significance level was set at 0.05.

## Results

### Patient Demographic Characteristics

For the primary cohort, among 182 patients with pathologically confirmed GBM, we excluded 16 patients due to unknown MGMT methylation results, 18 patients whose IDH were mutant, as well as 7 patients with missing images or poor image quality or failed format conversion. For the external validation cohort, 25 patients with pathologically confirmed GBM were included, with 10 patients excluded due to the absence of images or poor image quality or failed format conversion or unknown MGMT methylation. Finally, 156 patients were included, and the characteristics of patients in the training cohort (n = 113), internal verification cohort (n = 28), and external verification cohort (n = 15) were summarized as shown in Figure S1 and Table 1.

### Analysis of MGMT-Related Deep Learning Radiopathomics Features

Through correlation analysis between DLRP features and MGMT genotyping, a classification model for predicting MGMT genotyping was developed using integrated deep learning features from radiology images and pathology images. After ranking the Pearson correlation coefficient between deep learning radiopathomics features and labels in the training cohort, the top 128 features were initially selected. Subsequent to the application of the LASSO algorithm, features with non-zero weights were excluded, resulting in a total of 28 features employed for the construction of the DLRPM. The feature set used for model construction includes 11 DLR features and 17 DLP features. The specific features and their LR coefficients are listed in Supplementary Table S1. For comparison, the DLPM and DLRM also use the same construction method. The DLRM initially selected 64 features, and 11 DLR features were retained after LASSO analysis. DLPM initially retained 160 features, and LASSO analysis retained 29 DLP features. The feature selection results of building these two models are provided in the Supplementary Table S2 and Table S3.

### Assessment of Diagnostic Efficacy of Models for Predicting MGMT

The results of the study showed that the AUCs for predicting MGMT methylation status in the training and internal validation cohorts were 0.786 (95% CI 0.703–0.868), 0.771 (95% CI 0.586–0.956) for DLRM, 0.864 (95% CI 0.798–0.932), 0.844 (95% CI 0.691–0.996) for DLPM, 0.920 (95% CI 0.870–0.968), and 0.854 (95% CI 0.702–1) for DLRPM. DLRPM has significantly improved the prediction performance. The results of DLRPM are displayed in Figure 3a and Table 2. The results of DLRM and DLPM are displayed in Supplementary Figure S2-3 and Table 2.

**Figure 3.**
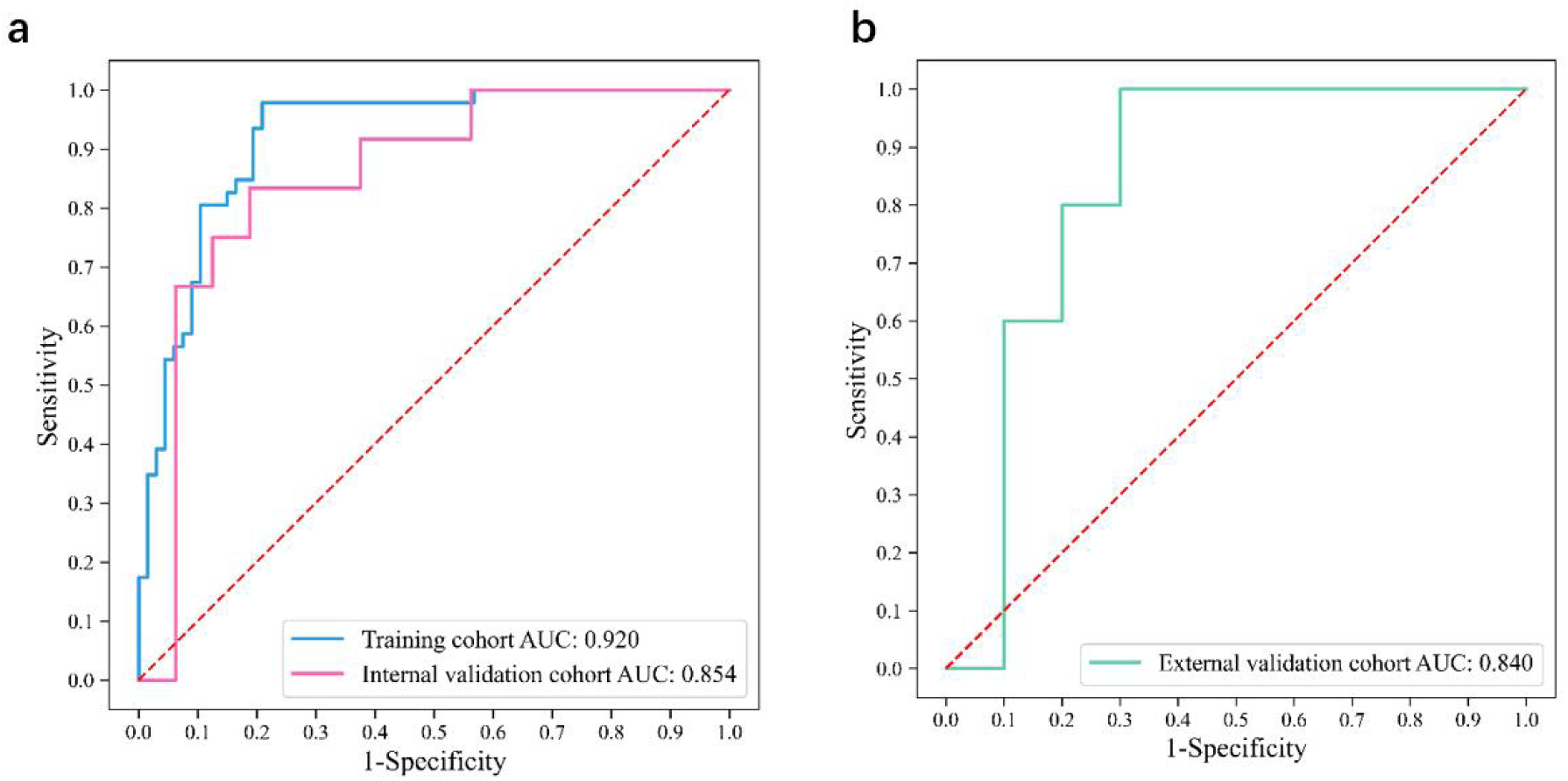
ROC curves show the ability of the Deep Learning Radiologic Pathology Model (DLRPM) to distinguish between the MGMT methylation states of the training cohort and the internal (a) and external validation cohorts (b). The red dashed line represents the diagonal.

**Table 2.** Area under the ROC curve (AUC), 95% confidence interval (CI), accuracy, sensitivity, and specificity of the DLRM, DLPM, and DLRPM for the training and internal validation cohorts and external validation cohorts.

| Cohort | Method | AUC (95% CI) | Accuracy | Sensitivity | Specificity |
| --- | --- | --- | --- | --- | --- |
| Training cohort | DLRM | 0.786 (0.703–0.868) | 0.673 | 0.804 | 0.582 |
|  | DLPM | 0.864 (0.798–0.932) | 0.796 | 0.717 | 0.851 |
|  | DLRPM | 0.920 (0.870–0.968) | 0.832 | 0.804 | 0.851 |
| Internal validation cohort | DLRM | 0.771 (0.586–0.956) | 0.679 | 0.750 | 0.625 |
|  | DLPM | 0.844 (0.691–0.996) | 0.786 | 0.600 | 0.875 |
|  | DLRPM | 0.854 (0.702–1) | 0.821 | 0.833 | 0.813 |
| External validation cohort | DLRM | 0.600 (0.292–0.908) | 0.600 | 0.800 | 0.500 |
|  | DLPM | 0.780 (0.695–1) | 0.667 | 0.800 | 0.600 |
|  | DLRPM | 0.840 (0.625–1) | 0.800 | 0.800 | 0.800 |

Abbreviations: DLRM, deep learning radiomics model; DLPM, deep learning pathomics model; DLRPM, deep learning radiopathomics model; AUC, area under the receiver operating characteristic curve.

### Model Predictions Generalization to Independent Validation Cohorts

We validated the models in an independent external validation cohort. The results of the study showed that the AUCs for predicting MGMT methylation status in the external validation cohort were 0.600 (95% CI 0.292–0.908) for DLRM, 0.780 (95% CI 0.695–1) for DLPM, 0.840 (95% CI 0.625–1) for DLRPM. DLRPM also has significantly improved the prediction performance. The results of DLRPM are displayed in Figure 3b and Table 2. The results of DLRM and DLPM are displayed in Supplementary Figure S4-5 and Table 2.

The model calibration curves are shown in Figure 4a, which were used to assess the degree of agreement between the model predictions and the actual situation. In the primary and external validation datasets, the calibration curves predicted by DLRPM were closest to the diagonal line compared to DLRM and DLPM, indicating that the model-predicted MGMT methylation status was highly consistent with the actual clinical situation, and showed a high degree of reliability and accuracy. Similarly, the decision curves showed the net benefit for DLRM, DLPM, and DLRPM over a range of threshold probabilities, with the DLRPM showing the largest net benefit, as shown in Figure 4b.

**Figure 4.**
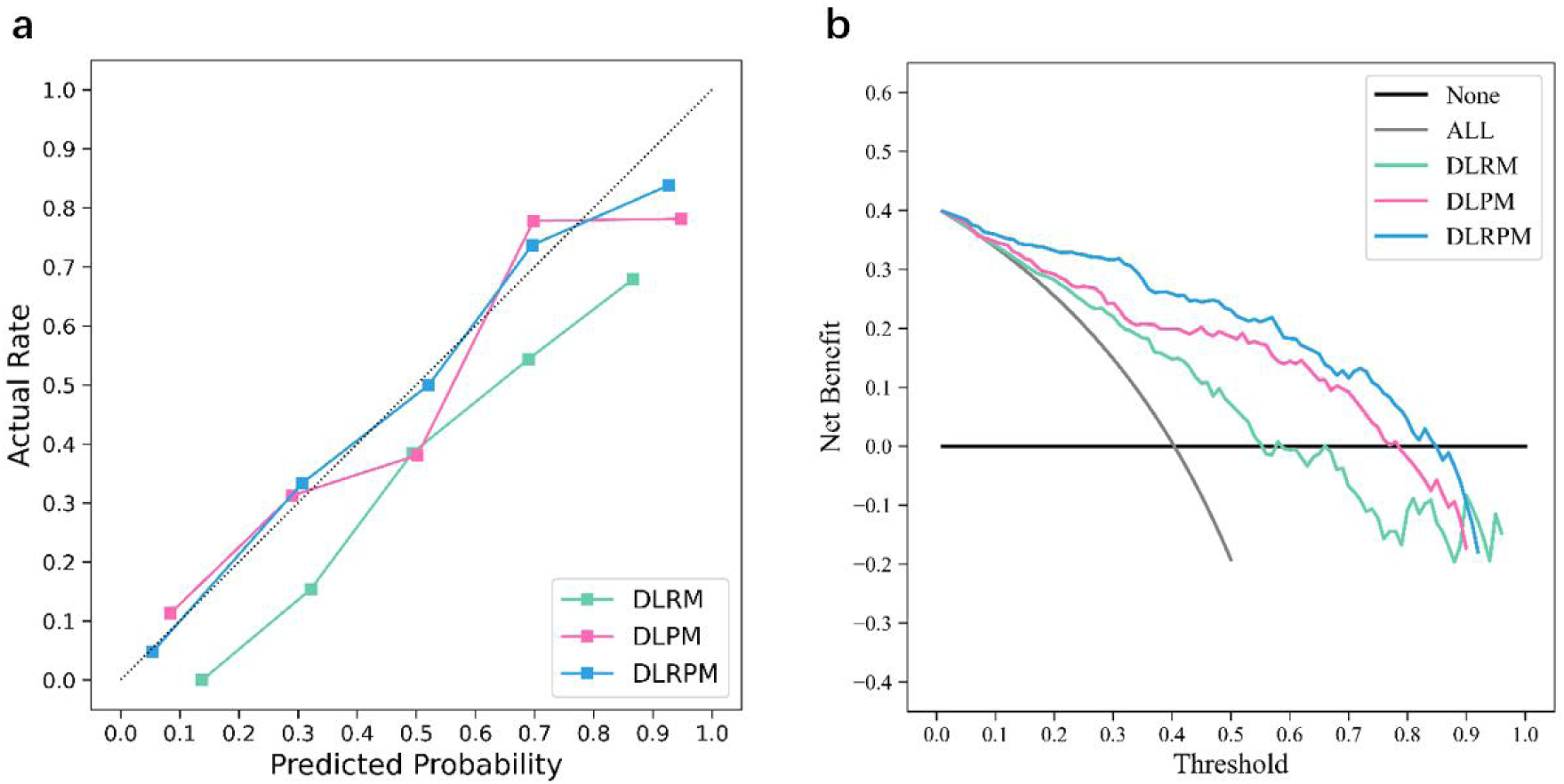
Model prediction performance. (a) The model calibration curve of the deep learning radiomics model (DLRM), deep learning pathology model (DLPM), and deep learning radiomics pathology model (DLRPM). The horizontal axis represents the predicted probability, the vertical axis represents the actual probability, and the diagonal line (black dashed line) is the ideal calibration curve. (b) Decision curve analysis shows that DLRPM has the highest net benefit compared to DLRM, DLPM. All, interfere with everyone. None, not interfere with anyone.

### Correlation between Model Predicted Values and Visualized MRI Features

Furthermore, we analyzed whether DLRPM was associated with lesion location, cystic necrosis, enhancement pattern, subventricular zone involvement, peritumoral edema, and lesion number. The results demonstrated that the predicted values in the MGMT-positive group showed significant correlation with subventricular zone involvement (P < 0.05). These findings are presented in Table 3.

**Table 3.** Spearman correlation between DLRPM predicted values and visualized MRI features.

|  | MGMT Methylation<br>(n=93) | MGMT Unmethylated<br>(n=63) |
| --- | --- | --- |
| Lesion location | $r = .158, P = .216$ | $r = .050, P = .632$ |
| Cystic necrosis | $r = -.005, P = .967$ | $r = .056, P = .597$ |
| Strengthening mode | $r = .063, P = .624$ | $r = -.022, P = .837$ |
| Subventricular zone | $r = .332, P = .008^*$ | $r = -.164, P = .115$ |
| Edema | $r = -.026, P = .839$ | $r = .095, P = .364$ |
| Number of lesions | $r = -.020, P = .877$ | $r = -.103, P = .326$ |

Note: \* Indicates statistical significance.

## Discussion

Predicting MGMT methylation status in GBM patients based on preoperative MRI and postoperative pathological sections has the potential to replace molecular detection and facilitate individualized and convenient clinical diagnosis and treatment decisions. Therefore, in this study, the AUC of DLRPM constructed by analyzing 156 cases of multiple whole brain MRI sequences (T1- and T2-weighted, T1-enhanced) and WSI-based whole pathology images were superior to those of DLRM and DLPM in both the training cohort, internal and external validation cohorts. To the best of our knowledge, this is the first time to predict MGMT methylation status in GBM patients based on deep learning combined with macro radiology and microscopic pathology. In addition, we used DCA to evaluate the net benefit of the three models, and the results showed that DLRPM had the highest net benefit, which means that we can recommend DLRPM to help determine MGMT methylation status. Our study provides a reliable and reproducible tool to predict the MGMT methylation status. Patients with MGMT methylation have a better prognosis and are more likely to benefit from adjuvant therapy (Flies et al., 2024).

While previous molecular prediction studies based on classical radiomics achieved high AUCs in internal validation in a single-center setting (Xi et al., 2018; Doniselli et al., 2023), DLRPM in this study was able to show similar performance, and was validated in a larger sample size and independent external cohort, and significantly outperformed the conventional single-scale prediction models. In addition, traditional radiomics feature extraction requires precise manual outlining of target areas, which limits the reproducibility and scalability of the study. Deep learning techniques have been successfully used for the prediction of other clinically relevant features of gliomas (Li et al., 2022) such as IDH, 1p/19q, and tumor grading (van der Voort et al., 2023). In this study, we use whole brain images as input based on a deep learning approach, which can significantly reduce the workload of subsequent studies and improve the comprehensiveness of features. In addition, fusion features of multiple MRI sequences with pathological tissue are a major highlight of this study, and integrating features from different dimensions into the model is challenging. We used deep learning as our radiological and pathological feature extractors, respectively, and integrated these two feature sets by the arithmetic mean after normalization to achieve a preliminary cross-scale study.

MGMT is a gene encoding a DNA repair protein that is critical for genomic stability. It is widely accepted that the methylation status of the MGMT promoter is a favorable prognostic factor for patients with GBM (Flies et al., 2024), and is associated with a stronger response to alkylating agents such as temozolomide, a higher response to radiotherapy, and a longer survival time (Hegi et al., 2005; Silva et al., 2024). However, molecular testing is associated with high costs, delayed results, and requires specialized laboratory equipment and trained personnel for operation. We combine radiology and pathology with the aim of improving prediction accuracy, convenience, and robustness, and introducing new insights into cross-scale information integration. Similarly, Feng et al. (Feng et al., 2022) found that an integrated radiopathomics prognostic system was able to predict the pathological complete response of rectal cancer to neoadjuvant radiotherapy based on preprocessed radiopathology images with high accuracy and robustness. Sun et al. (Sun et al., 2021) found that prognostic radiomics MRI phenotypes of GBM patients are associated with different biological pathways through radiogenomics. Deep learning radiomics involves high-throughput extraction of computational features that quantify tissue heterogeneity at the macroscopic level using advanced image processing and computer vision techniques. Whereas deep learning pathomics provides quantitative information at the microscopic level, providing methods for implementing personalized and precise treatment and prognosis prediction (Volinsky-Fremond et al., 2024b). Fusion of deep learning radiomics and pathomics features provides an opportunity to combine tumor heterogeneity at the macro and micro scale, which may complement each other and result in a stronger integrated signature (Lu et al., 2021a).

Recent research suggests that if the goal is to guide decision making through prediction, the clinical utility of predictive models should also be assessed, e.g., using net benefits and decision curves (Riley et al., 2024). DCA is a statistical technique for evaluating tests or models focusing on decisions and outcomes (Vickers and Woo, 2022), and DCA provides us with a clear answer about whether the benefits of a model outweigh its drawbacks. This contrasts with traditional measures such as sensitivity, specificity, or area under the curve, which are abstract statistics that do not directly provide information of clinical value. In our DCA study, the net benefit of DLRPM was higher than that of both DLRM and DLPM, and was positive over a wider range of thresholds. This means that we can recommend the model to help determine the methylation status of MGMT.

Our study has several limitations. First, all data were retrospectively collected, and both internal and external validation cohorts had relatively small sample sizes, potentially introducing selection bias that may have influenced our results. Second, the lack of precise information regarding surgical specimen orientation and location precluded further exploration of the correlation between radiological and pathological features. Third, long-term survival follow-up data were unavailable. Additionally, while deep learning models function as “black boxes” that autonomously extract features without human-defined morphological assessments, the primary challenge lies in their limited interpretability. However, we are actively working to address this through visualization techniques and quantitative analyses to elucidate the imaging and pathological features that the model focuses on. Therefore, validation with larger prospective cohorts is warranted. Despite these limitations, our results demonstrate that DLRPM maintained its performance in independent external validation, even with variations in MRI scanning parameters and pathological staining protocols across different cohorts. This underscores the robust capability of DLRPM in detecting MGMT promoter methylation status.

In conclusion, we developed and validated a deep learning radiopathomics model based on preoperative MRI and postoperative pathological tissues that predicts the genotype of GBM patients without any manual feature extraction or selection. The performance and generalizability of this model highlights the potential for application in adjunctive treatment decision-making for IDH wild-type GBM patients. In the future, more patient groups should be included to further validate and optimize the model performance. Cross-scale omics integration research may represent the direction of future cancer research.

## Data Availability

All data produced in the present study are available upon reasonable request to the authors

## Author contributions

Guarantors of integrity of entire study, X.W., W.W., H.Y., Y.B.X.; study concepts/study design or data acquisition or data analysis/interpretation, all authors; manuscript drafting, X.W., M.M.; manuscript revision for important intellectual content, all authors; approval of final version of submitted manuscript, all authors; agrees to ensure any questions related to the work are appropriately resolved, all authors; literature research, X.W., M.M.; clinical studies, X.W., Y.L., Y.Q.X., W.H., G.C., X.K., Y.B.X., H.Y.; experimental studies, X.W., W.W., Y.L., M.M., Y.B.X.; statistical analysis, X.W., M.M.; and manuscript editing, all authors.

## Acknowledgements

The authors extend their sincere appreciation to Dr. Mengchen Hou from the Department of Pathology and Dr. Tengyu Guo from the Department of Radiology at Xi’an People’s Hospital (Xi’an Fourth Hospital) for their invaluable insights and professional expertise, which have made significant contributions to this research.

## Availability of data and materials

The data that support the findings of this study are available from the corresponding author, Yibin Xi, upon reasonable request.

## Declarations

### Ethical approval

The Institutional Review Board of Xi’an People’s Hospital (Xi’an Fourth Hospital) approved the study protocol (No. 230306039), and the study was conducted in accord with the Helsinki Declaration.

### Consent to participate

Due to this being a retrospective study, informed consent was waived by the institutional review board.

