## Supplementary Table S1 for "Cross-scale prediction of glioblastoma MGMT methylation status based on deep learning combined with magnetic resonance images and pathology images"

**Running title: Radiopathomics-based MGMT methylation prediction**

**ABSTRACT:**

**Background:** In glioblastoma (GBM), promoter methylation of the O^6^-methylguanine-DNA methyltransferase (MGMT) is associated with beneficial chemotherapy but has not been accurately evaluated based on radiological and pathological sections. To develop and validate an MRI and pathology image-based deep learning radiopathomics model for predicting MGMT promoter methylation in patients with GBM.

**Methods:** A retrospective collection of pathologically confirmed isocitrate dehydrogenase (IDH) wild-type GBM patients (n=207) from three centers was performed, all of whom underwent MRI scanning within 2 weeks prior to surgery. The pre-trained ResNet50 was used as the feature extractor. Features of 1024 dimensions were extracted from MRI and pathological images, respectively, and the features were screened for modeling. Then feature fusion was performed by calculating the normalized multimode MRI fusion features and pathological features, and prediction models of MGMT based on deep learning radiomics, pathomics, and radiopathomics (DLRM, DLPM, DLRPM) were constructed and applied to internal and external validation cohorts.

**Results:** In the training, internal and external validation cohorts, the DLRPM further improved the predictive performance, with a significantly better predictive performance than the DLRM and DLPM, with AUCs of 0.920 (95% CI 0.870–0.968), 0.854 (95% CI 0.702–1), and 0.840 (95% CI 0.625–1).

**Conclusion:** We developed and validated cross-scale radiology and pathology models for predicting MGMT methylation status, with DLRPM predicting the best performance, and this cross-scale approach paves the way for further research and clinical applications in the future.

**KEYWORDS:** Glioblastoma, Deep learning, MRI, Radiopathomics, Genotypes

**Supplementary materials**

**Methods:**

The scan sequences of primary dataset included the following: (1) axial T1WI: TR, 250 ms; TE, 2.5 ms; section thickness, 5 mm; inter-slice gap, 0 mm; field of view (FOV), 240 mm × 240 mm; matrix, 230 × 256; (2) axial T2WI: TR, 4000 ms; TE, 91 ms; section thickness, 5 mm; FOV, 240 mm × 240 mm; matrix, 320 × 320; (3) axial post-contrast T1 was acquired after intravenous administration of 0.1 mmol/kg gadopentetate dimeglumine. The external validation dataset including axial T1WI (TR, 250 ms; TE, 2.5 ms; section thickness, 5 mm; matrix, 320 × 256), axial T2WI (TR, 6000 ms; TE, 96 ms; section thickness, 5 mm; matrix, 320 × 320), and axial post-contrast T1.

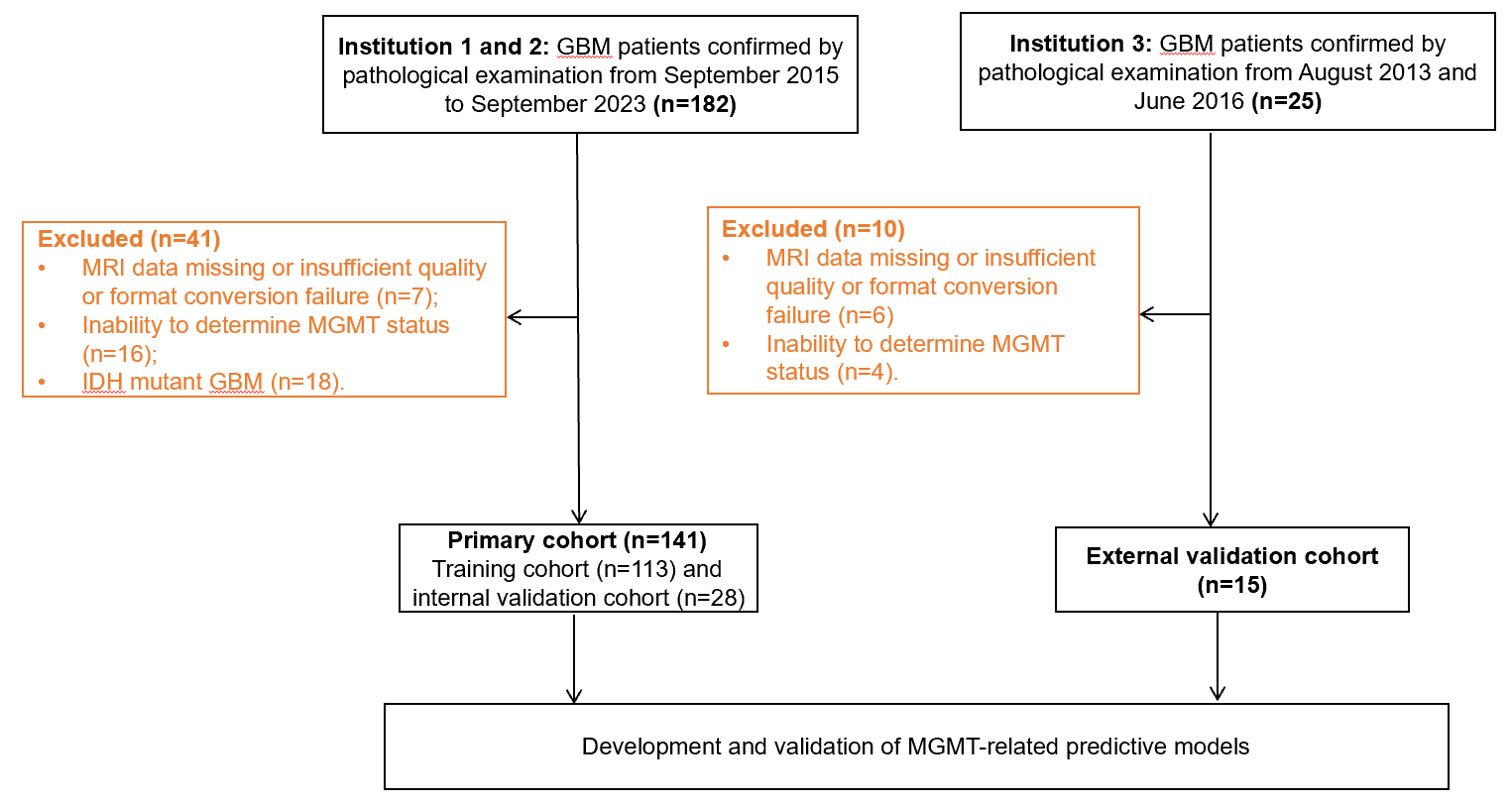

**Figure S1.** Flowchart showing glioblastoma (GBM) patient enrollment, including the training and internal validation cohorts and the external validation cohort.

**Results:**

**Table S1: Feature Selection Outcomes for DLRPM Construction**

| Feature origin | Feature name | Coefficients |
| --- | --- | --- |
| MRI  (N=11) | DLR_234 | -0.286365163 |
|  | DLR_326 | 0.074104483 |
|  | DLR_488 | -1.597194045 |
|  | DLR_508 | -2.111834323 |
|  | DLR_516 | 1.410934019 |
|  | DLR_547 | 1.489454829 |
|  | DLR_562 | -0.970416642 |
|  | DLR_813 | 0.743132769 |
|  | DLR_837 | 0.950954352 |
|  | DLR_896 | -0.738256362 |
|  | DLR_1000 | -1.968256185 |
| WSI  (N=17) | DLP_16 | -0.295561132 |
|  | DLP_31 | 0.510422498 |
|  | DLP_33 | -0.964813688 |
|  | DLP_39 | 1.284574874 |
|  | DLP_87 | -0.201792466 |
|  | DLP_91 | -0.37527694 |
|  | DLP_98 | -0.859611241 |
|  | DLP_113 | -1.463793262 |
|  | DLP_120 | 1.166544724 |
|  | DLP_194 | 1.695869068 |
|  | DLP_209 | 0.339146103 |
|  | DLP_235 | 0.224612067 |
|  | DLP_284 | 0.455664043 |
|  | DLP_446 | -1.730384155 |
|  | DLP_508 | 1.209909375 |
|  | DLP_529 | -0.073132363 |
|  | DLP_697 | 0.500259522 |

Note: Feature analysis conducted using Pearson correlation and LASSO algorithm. The coefficients represent the magnitude of feature weights in the DLRPM.

**Table S2: Feature Selection Outcomes for DLRM Construction**

| Feature origin | Feature name | Coefficients |
| --- | --- | --- |
| MRI  (N=11) | DLR_73 | 0.783933072 |
|  | DLR_234 | -0.464476455 |
|  | DLR_369 | -0.451266548 |
|  | DLR_488 | -0.609749859 |
|  | DLR_508 | -0.902913065 |
|  | DLR_516 | 0.207732037 |
|  | DLR_547 | 0.789957522 |
|  | DLR_562 | -0.767006836 |
|  | DLR_813 | 0.125759587 |
|  | DLR_896 | -0.416450513 |
|  | DLR_1000 | -1.342830771 |

**Table S3: Feature Selection Outcomes for DLPM Construction**

| Feature origin | Feature name | Coefficients |
| --- | --- | --- |
| WSI  (N=29) | DLP_16 | 0.671965305 |
|  | DLP_33 | -0.134014238 |
|  | DLP_39 | 0.895988851 |
|  | DLP_56 | 0.158493884 |
|  | DLP_71 | 0.390335895 |
|  | DLP_87 | 0.259198349 |
|  | DLP_91 | -0.486638913 |
|  | DLP_98 | -1.198991212 |
|  | DLP_113 | -0.783756048 |
|  | DLP_235 | -0.450646928 |
|  | DLP_284 | 1.392391534 |
|  | DLP_353 | 0.301548606 |
|  | DLP_386 | 1.08221846 |
|  | DLP_390 | 0.219354997 |
|  | DLP_446 | -2.160337911 |
|  | DLP_508 | 0.703360073 |
|  | DLP_529 | 0.135136555 |
|  | DLP_557 | 1.260092594 |
|  | DLP_573 | -0.323991293 |
|  | DLP_594 | 0.154988125 |
|  | DLP_683 | 1.630342916 |
|  | DLP_697 | 0.19820991 |
|  | DLP_758 | 0.246712892 |
|  | DLP_781 | -1.131343178 |
|  | DLP_889 | 1.10589376 |
|  | DLP_950 | 0.643299652 |
|  | DLP_953 | -0.312445462 |
|  | DLP_1014 | 0.196230291 |
|  | DLP_1015 | -0.248813086 |

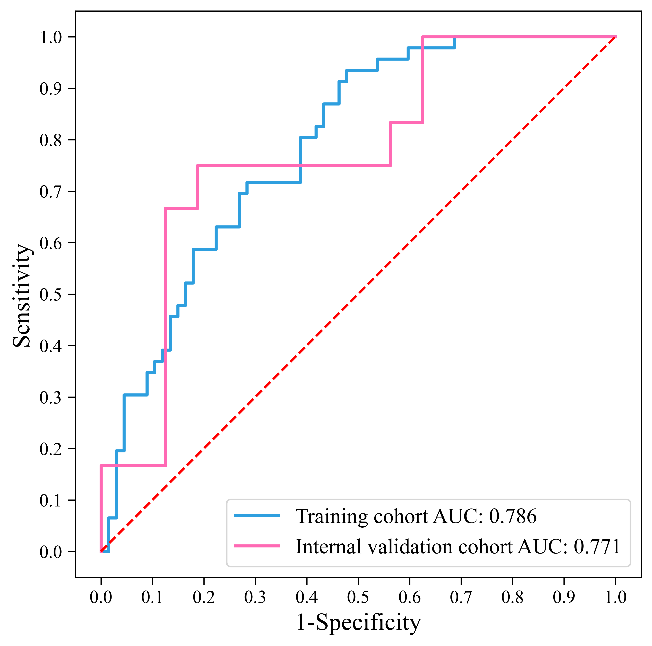

**Figure S2.** The ROC curve shows the ability of the deep learning radiomics model (DLRM) to distinguish MGMT methylation status with significance. The area under the ROC curve in the training cohort (blue line) was 0.786, and the area under the ROC curve in the internal validation cohort (red line) was 0.771. The red dashed line represents the diagonal line.

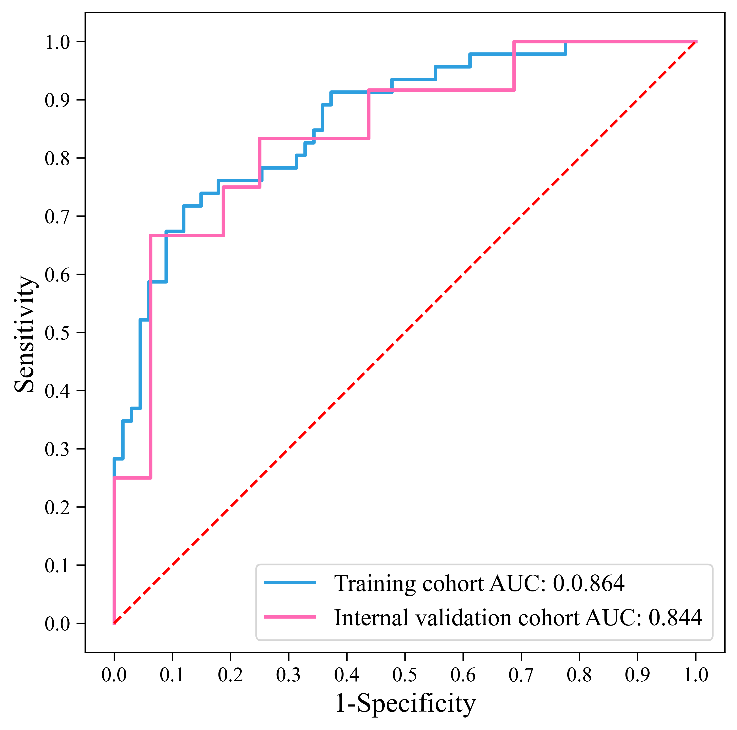

**Figure S3.** The ROC curve shows the ability of the deep learning pathomics model (DLPM) to distinguish MGMT methylation status with significance. The area under the ROC curve in the training cohort (blue line) was 0.864, and the area under the ROC curve in the internal validation cohort (red line) was 0.844. The red dashed line represents the diagonal line.

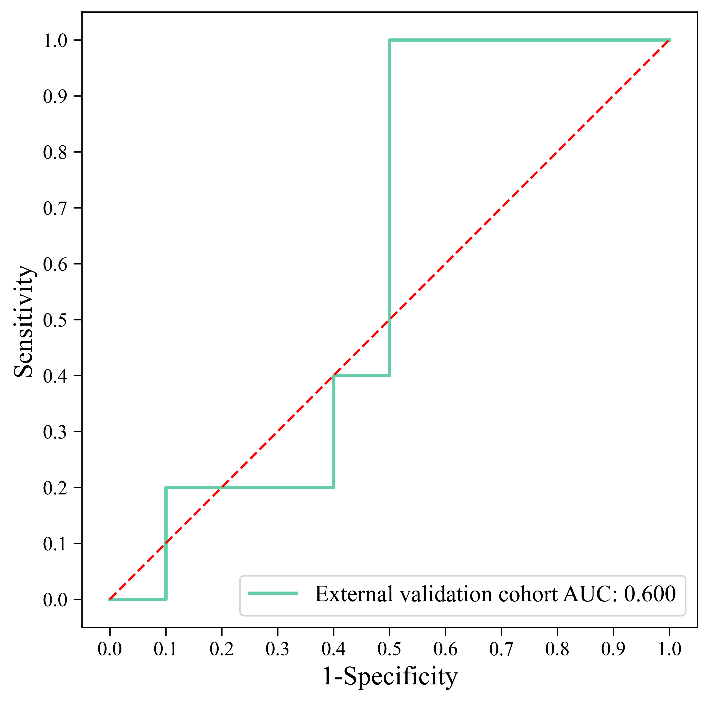

**Figure S4.** The ROC curve shows the ability of the deep learning radiomics model (DLRM) to distinguish MGMT methylation status with significance. The area under the ROC curve in the external validation cohort (green line) was 0.600. The red dashed line represents the diagonal line.

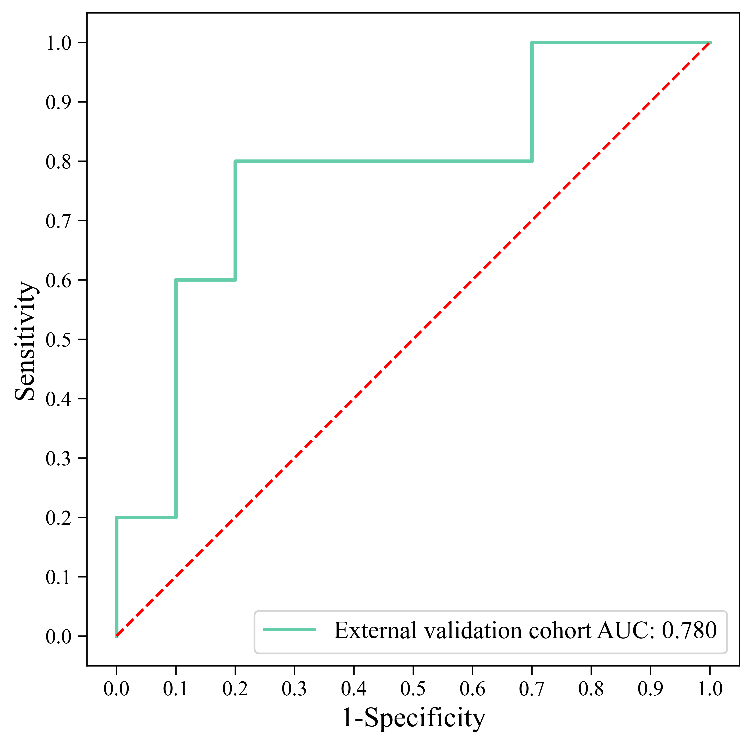

**Figure S5.** The ROC curve shows the ability of the deep learning pathomics model (DLPM) to distinguish MGMT methylation status with significance. The area under the ROC curve in the external validation cohort (green line) was 0.780. The red dashed line represents the diagonal line.
